## Supplementary File 1 for "Using Digital Tools in Clinical, Health and Social Care Research: A Mixed-Methods Study of UK Stakeholders"

**Supplementary Material**

**Table S1:** Definition of digital tool by survey type

| Research Staff | Research and Development |
|---|---|
| In this study a **digital tool** is defined as an alternative to paper-based methods that is IT based or an online platform that aids any aspect of the research study (stages of clinical research include but are not limited to; set up of research studies, participant recruitment, consent, participant retention, intervention delivery, data collection, data analysis, monitoring, patient reported outcomes).<br>Examples of digital tools might include but are not limited to:<br>• Electronic databases for participant screening<br>• Social media platforms for participant recruitment<br>• Communication within your research team (e.g., reporting adverse events via text message or email)<br>• Electronic consent<br>• Text messaging for participant retention<br>• Digital platforms for collecting survey data | In this study a **digital tool** is defined as an alternative to paper-based methods that is IT based or an online platform that aids any aspect of the research study set up, recruitment of participants and management.<br>Examples of digital tools might include but are not limited to:<br>• Electronic screening for participants at site level<br>• Remote monitoring via smart phone applications<br>• Local social media routes<br>• R&D department’s website<br>• Text alerts |

**Table S2:** Summary of Question area by survey type (Research Staff, R&D)

| Survey Type | Question Themes |
|---|---|
| **Research Staff** | **Most effective digital tool** |
| | · Description of tool |
| | · Type of research digital tool used within (e.g., clinical trial) |
| | · Stage of research project tool related to (e.g., recruitment) |
| | · Project status |
| | · Widely used/bespoke tool |
| | · Costs associated with acquiring tool |
| | · Training required to use tool |
| | · End user of tool (drop down menu of pre-specified answers) |
| | · Effectiveness of the digital tool relative to other digital tools (0 = not effective, 10 = very effective) |
| | · How essential was digital tool to successfully implement the research (relative to alternative non-digital tools) (1 Highly non-essential to 5 Highly essential) |
| | · Effectiveness of integration of digital tool with other health care or research systems (1 Not applicable to 5 highly effective) |
| | · Level of technical literacy required to use/access tool (1 Very difficult to use/access to 4 Very easy to use/access) |
| | · Main goal/s of tool (drop down menu of pre-specified answers) |
| | Least effective digital tool |
| | · Same as above except for Ineffectiveness of the digital tool relative to other digital tools 0 = no less effective than other tools, 10 = much less effective than other tools. |
| **R&D** | **Most effective digital tool example for set up** |
| | · Description of tool |
| | · How heard about digital tool |
| | · Training (if yes number of days) |
| | · Widely used/bespoke novel tool |
| | · Cost information |
| | · Perceived cost effectiveness of tool |
| | Most effective digital tool example for recruitment |
| | · Same as above |
| | Most effective digital tool example for management |

| | |
|---|---|
| | · Same as above |

**Table S3:** Summary of interview question areas by participant type

| Interview Type | Question Themes |
|---|---|
| Researchers and Related Staff | Definition of digital tools<br>How did COVID-19 affect role/use of digital tools<br>Case study about most effective tool<br>Case study about least effective tool<br>Any instances where tools could be used<br>Any barriers to digital tools<br>What would help researchers when deciding what tool to use/where to find this information |
| R&D Staff | Definition of digital tools<br>How did COVID-19 affect role/use of digital tools<br>Case study most effective tool for set up<br>Case study most effective tool for recruitment<br>Case study most effective tool for management<br>Any instances where tools could be used<br>Any barriers to digital tools<br>What would help researchers when deciding what tool to use/where to find this information |

**Table S4:** Research staff and related survey participant characteristics

| Number of Participants (N=41) | N (%) |
|---|---|
| *County* | |
| County of the West Midlands | 26 (63.4) |
| Warwickshire | 9 (22) |
| Staffordshire | 4 (9.8) |
| Multiple counties in the CRN West Midlands | 2 (4.9) |

| | |
|---|---|
| *Employing organisation* | |
| University | 14 (34.1) |
| Clinical Trials Unit | 13 (31.7) |
| NHS Trust | 11 (26.8) |
| Other (please provide further details below) | 3 (7.3) |
| *Job role* | |
| Chief Investigator | 8 (19.5) |
| Other | 8 (19.5) |
| Trial/Study Coordinator | 8 (19.5) |
| Trial/Study Manager | 8 (19.5) |
| Research Manager | 3 (7.3) |
| Principal Investigator | 2 (4.9) |
| Research Nurse | 2 (4.9) |
| Trial/Study Data Manager | 2 (4.9) |

**Table S5** Most effective and least effective digital tools results

| Question Type | Answer Options | Most Effective N(%) | Least Effective N (%) |
|---|---|---|---|
| Type of research project | Clinical Trial | 8 (38.1) | 9 (42.9) |
| | Clinical Research | 6 (28.6) | 6 (28.6) |
| | Health Research | 5 (23.8) | 4 (19) |
| | Other | 1 (4.8) | 1 (4.8) |
| | Social Care | 1 (4.8) | 1 (4.8) |
| | Total number of responses | 21 | 21 |
| Stage of research | Research study set up | 2 (9.5) | 4 (19) |
| | Participant recruitment | 4 (19) | 8 (38.1) |
| | Data collection | 11 (52.4) | 6 (28.6) |
| | Intervention delivery | 2 (9.5) | 1 (4.8) |
| | Quality Assurance | n/a | 1 (4.8) |
| | Outcome measures | 1 (4.8) | n/a |

| | | | |
|---|---|---|---|
| | Other | 1 (4.8) | 1 (4.8) |
| | Total number of responses | 21 | 21 |
| Project status | Completed | 4 (19) | 4 (19) |
| | Ongoing | 17 (81) | 17 (81) |
| | Total number of responses | 21 | 21 |
| Widely used vs Bespoke digital tool | Novel/bespoke to my organisation | 4 (19) | 3 (14.3) |
| | Novel/bespoke to my project team | 5 (23.8) | 1 (4.8) |
| | Used widely | 12 (57.1) | 17 (81) |
| | Total number of responses | 21 | 21 |
| Costs associated with acquiring digital tool | A one-off up-front fee | 1 (4.8) | 1 (4.8) |
| | A one-off up-front fee, periodic recurring fee | 2 (9.5) | 1 (4.8) |
| | I'm not sure | 4 (19) | 8 (38.1) |
| | No fee (e.g., open source) | 8 (38.1) | 7 (33.3) |
| | Other | 3 (14.3) | 3 (14.3) |
| | Periodic recurring fee | 3 (14.3) | 1 (4.8) |
| | Total number of responses | 21 | 21 |
| Training required in order to use tool | No | 11 (52.4) | 12 (57.1) |
| | Yes | 10 (47.6) | 9 (42.9) |
| | Total number of responses | 21 | 21 |
| How essential was this tool to successfully implement the research | Somewhat non-essential (2) | 1 (4.8) | 4 (19) |
| | Somewhat essential (3) | 8 (38.1) | 9 (42.9) |
| | Highly essential (4) | 12 (57.1) | 8 (38.1) |
| | Total number of responses | 21 | 21 |
| Integration with other healthcare or research systems | Not applicable (1) | 3 (14.3) | 2 (9.5) |
| | Somewhat ineffective (3) | 3 (14.3) | 6 (28.6) |
| | Somewhat effective (4) | 8 (38.1) | 10 (47.6) |
| | Highly effective (5) | 7 (33.3) | 3 (14.3) |
| | Total number of responses | 21 | 21 |

| | | | |
|---|---|---|---|
| Level of technical literacy required | Somewhat difficult to use/access (2) | 1 (4.8) | 7 (33.3) |
| | Somewhat easy to use/access (3) | 10 (47.6) | 10 (47.6) |
| | Very easy to use/access (4) | 10 (47.6) | 4 (19) |
| | Total number of responses | 21 | 21 |

**Table S6:** Rating of effectiveness of most effective tool (0 no more effective to 10 much more effective)

| Answer Options | Rating of most effective tool compared to other tools |
|---|---|
| 0 | n/a |
| 1 | n/a |
| 2 | n/a |
| 3 | n/a |
| 4 | n/a |
| 5 | 1 (4.8%) |
| 6 | 1 (4.8%) |
| 7 | 4 (19%) |
| 8 | 7 (33.3%) |
| 9 | 5 (23.8%) |
| 10 | 3 (14.3%) |
| Total number of responses | 21 |

**Table S7:** Rating of ineffectiveness of least effective tool (0 = no less effective than other tools, 10 = much less effective than other tool)

| Answer Options | Rating of least effective tool compared to other tools |
|---|---|
| 0 | 1 (5%) |
| 1 | 1 (5%) |
| 2 | 2 (10%) |
| 3 | 2 (10%) |
| 4 | 3 (15%) |
| 5 | 7 (35%) |
| 6 | 1 (5%) |
| 7 | 2 (10%) |
| 8 | 1 (5%) |
| 9 | n/a |
| 10 | n/a |
| Total number of responses | 20 |

**Table S8:** R&D Survey Participant Characteristics

| Number of Participants (N=25) | N(%) |
|---|---|
| *Experience Using Digital Tools* | |
| Participants who used digital tools | 22(88) |
| Participants who have not used digital tools | 3(12) |
| *County* | |
| County of the West Midlands | 10(40) |
| Warwickshire | 5(20) |
| Multiple counties in CRN West Midlands | 3(12) |

| | |
|---|---|
| Staffordshire | 3(12) |
| Worcestershire | 2(8) |
| Herefordshire | 1(4) |
| Shropshire | 1(4) |
| *Employing Organisation* | |
| NHS | 25(100%) |
| *Job Role* | |
| R&D Manager/Head of R&D | 13(52) |
| Lead Research Nurse | 3(12) |
| Other (please provide further details below) | 3(12) |
| R&D Facilitator (governance, study set-up) | 3(12) |
| Data Manager | 1(4) |
| Research Delivery staff | 1(4) |

**Table S9:** Most effective tool case studies for management and recruitment

| **Case study 1: Digital Tool for Management** | **Example Statements** |
|---|---|
| Two participants referred to a bespoke management tool as their most effective digital tool. The tool was designed and developed by R&D staff supported by a digital team embedded in an R&D department in collaboration with clinical research staff (primarily nurses). Built in excel, the tool is used to manage patients through a study protocol. Below, one participant describes the tool. | *"It meets multiple needs, so we build them individually off the back of each protocol…so when you put the patients recruited date it then predicts all their follow up visits for you in keeping with the protocol. It then has a traffic light system which would tell you when they're due to come up so the teams can see it and have oversight quite quickly of all the participants where they are, what happens at each individual stage. You then hover your mouse over visit 2 and a little info box will come up and tell you a brief synopsis of what needs to be done at that visit. So going quickly, either give you what needs to be done, you can click on it, and it takes you through to the worksheets that we create off the back of it."* |
| One participant explained the primary reason for developing the tool was to standardise study management. | *"It came around a sort of a want to offer standardised high-quality research and oversight... I've worked in lots of different research areas, and they've always got slightly different way of working. Some of the nurses and the clinical teams have sort of had to go at building their own Excel spreadsheets. Some of the sponsors and the studies provided them for us and then some used paper diaries. So, but when you look at it, none of them were quite really fulfilling everyone's needs, there were sort of doing a bit here and a bit there and I wanted to standardize it across the whole of R&D because it wasn't quite equitable for our patients and our staff. Some were getting access to. So that's why we standardized and went we're gonna build these and offer them out across the board."* |
| Prior to development of this bespoke management tool, it appears that studies were managed via paper means and/or nurses had attempted to build their own databases to manage studies. | *"Nurses that trying to build databases, you know, it's not our skill set, but some can do sort of just a small amount with them and then others were very much using paper which is not the way to do it."* |
| Both participants deemed the bespoke management tool to be effective for a number of different reasons including helping staff keep patients to the protocol time frames. One participant explained that the tool can be tweaked to suit needs of the users (i.e., nurses) and new features can be added. They also explained that it has many benefits due to being developed in excel, including tool template can be sent to teams located at different sites and used without the R&D staff visiting that particular site as well as being user friendly. | *"In terms of what it gains and what it offers for clinical deliveries…if you use [tool name] your patient should stay within [the] protocol visit window…and you can plan a lot easier, and again should someone go off…someone can come in and pick it up quite easily and you can ensure the patient is safe and kept up to date."*<br><br>*"One of the benefits of it is we've built it in Excel. It doesn't look like excel when you use it. Everyone's got excel within the trust, and there's no additional cost apart from the hours to build it, we weren't having to purchase another system. So that was why we chose that setup to build it. And then we're working with other sites and helping them have access. We can send them a template, we don't need to go there…they upload the template, then they can use it themselves. So that's why we built it that way just for that reason."*<br>*"I think we're on version seven or eight now, so they're really user friendly and they're really locked down so you can't break them. I've had some nurses do some really weird things and they've broken [it] but generally as times gone by, we've ironed out all the things. Before if a nurse dragged a formula* |

| | |
|---|---|
| | *away and deletes it, it would affect everything else…but in the department we've [become] more savvy at locking things down. Generally, …you should be able to click and use it.”* |
| A further benefit highlighted by participants is the fact that the tool was developed in collaboration with clinical research staff and has incorporated their ideas into the tool such as automatically generating reports opposed to looking through paper documents. | *“It's made by the people delivering research…I think together we’ve developed the service over the years and the tracker itself and that's what makes it better than an out-of-the-box bit of kit from it or third-party developer…one of the things we built in which was one of the ideas from a nurse, she had nearly a 1000 patients on a study but she just wanted to see who she needed to see this month, and rather than scrolling down the 1000 patients with a pen seeing “Who do I need to see?” She clicks the month button, and it will automatically generate your report in PDF and just say these are your patients for the month…I think that's really where the track of kind of stands out. It's just that bit of being able to organize and manage yourself and your team a lot more effectively and actually you don't want to spend time trawling through bits of paper, paper calendars and diaries.”* |

| **Case Study 2: Digital Tool for Recruitment** | **Example Statements [Participant Number]** |
|---|---|
| One participant described digital methods used in a clinical trial to recruit participants who meet the inclusion criteria of a study. Using data from the CPRD, researchers are able to know how many patients within each GP surgery meet an inclusion criteria for a study. Advantages of this include reduction in time and money spent on recruitment, as well as increases the likelihood of recruiting relevant participants. It is also beneficial for the patient, as the GP surgery may have relevant information on file that research studies can use (e.g., blood pressure). This is in turn also reduces the workload for the research team. | *“Practices sign up to CPRD [Clinical Practice Research Datalink] and they've got around 20% of practices across England. So CPRD are able to extract their coded medical data. So, any information that when you go to the GP and they say you've got diabetes, they put what they call a medical code in your patient records. So, you've got diabetes… CPRD able to extract that medical code...it's just that coded that this patient has diabetes, their blood pressure was this, they're on these drugs. They're able to extract that from practice records. So CPRD and [employing university] have spotted that actually there's a potential for using this in an interventional trial. So, it means there is an ability to extract that medical info, we can first of all pre-screen remotely the entire of CPRD's population, so over 13 million records we're able to pre-screen against our trial inclusion and exclusion criteria. So, we're then able to actually instead of the old-fashioned research way, which is you ask practices that do they want to take part in research? They say yes. Sounds interesting. You set them up, you spend time, money, setting them up, and actually they've got one patient…they haven’t got the patients you need. Whereas with this ability of CPRD’s, we're able to know that practice has this amount of patients…we're going to those practices that are large and potentially successful in actually taking part in research so that means obviously it reduces the time that we spend, the money that we spend and hopefully increases the success of the trial…and also obviously reduces practice workload... every so often a patient would go into the practice, have their blood pressure taken anyway for us then to ask that patient to come in for an extra research visit when actually we can just pull that data from the medical record, it seems counterintuitive, so making it easier one for practices they don't have to arrange for patients to come in and two for patients, it means they can absolutely take part in this trial without actually having to go into their GP practice.”* |

The participant explained that they also used e-consent in the trial when recruiting participant. Consent forms were sent to patients from the GP practises via text message or email. The participant explains the process below.

*“In response to COVID and the reduction in face-to-face GP visits we obviously needed to find a work around practices to be able to recruit patients to the trial. So, we actually developed a fully remote consent system using REDCap…we used it to send e-consent forms out to patients. So, the GP practice puts in some contact details of the patient and that then automatically sends a text message or e-mail to the patient containing a link to their consent form in this sort of electronic survey form. Patients then click on that link in that text message or e-mail it brings up their e-consent form. It provides them with the patient information sheet that they need to complete. Then they then go down and click yes, yes or no to each of the consent criteria. They then fill in their details. They're able to electronically sign that with that signature being saved so it matches that, that sort of the NHS criteria for e-consent system we then collect further information, so their e-mail and telephone number.*

The participant explains below the benefits of participants, GP and research team all receiving a copy of the e-consent form

*“and then once they've completed that it automatically notifies the GP practice that they've completed…the GP practice are then able to sign their section of the consent form and then it also automatically sends a copy of the consent form to the patient and to the practise. So, it automatically means that thinking about that good clinical practise the patients gets it, the practise gets it and us as a study team have that as a central repository so it makes it a lot easier for patient to consent… GP's obviously like that nature of being able to send the consent form out and then just being notified to say it's complete as well”.*

Other benefits include the ability to send out follow up questions in different languages.

*“We're able to use REDCap to automatically send out our follow up questionnaires. So, every six months we send quality of life questionnaires that again, patients are able to do on their phone, they're able to do in different languages. And they're able to then that would so complete that automatically comes back us and saves in the REDCap database… it's able to be used by patients and we've always had good response and patients have found it easy to complete and we get really good sort of follow up questionnaire response rate. I think we've got very few that haven't completed their follow up questionnaires”*
