## Supplementary File 2 for "Using Digital Tools in Clinical, Health and Social Care Research: A Mixed-Methods Study of UK Stakeholders"

### Supplementary Information 2a - Online Survey: Researchers and Related Staff

Start of Block: information sheet

Information sheet **PROJECT TITLE**: **NIHR Experiences of using Digital Tools in Clinical, Health and Social Care Research**

**NAMES OF RESEARCHERS:** Sophie Clohessy1, Prof Theo Arvanitis1, Dr Carla Toro1, Dr Mark Elliott1 Institute of Digital Health Care, WMG, University of Warwick)

**NAME OF COLLABORATORS:** Mark Evans2,3 Carly Craddock2,3 (2. The Royal Wolverhampton NHS Trust, Wolverhampton, UK, 3) National Institute for Health Research Clinical Research Network West Midlands, Birmingham, UK).

This sheet seeks to provide information, and advice, with respect to an individual's participation in support of the specified research project:

1. The study aims to investigate how online and digital tools/methods are being used for clinical, health and social care research. We are particularly interested to determine which approaches work well and which don't and use these findings to share best practice with the clinical, health and social care research community.

2. This study is funded by the National Institute of Health Research Clinical Research Network West Midlands (CRN West Midlands). The funding for this project has been awarded to Sophie Clohessy, Prof Theo Arvanitis, Dr Carla Toro, Dr Mark Elliott WMG, University of Warwick. The project will be led by The University of Warwick.

3. Participation in this research is totally voluntary, and assurances are given to the effect that no negative consequences will arise from refusal to participate in the research project.

4. Your consent for your data to be used in this questionnaire will be gained by your ticking the consent question at the end of this page, so by ticking this box you agree that your submitted data can be used in the aforementioned study.

5. Your data will be collected anonymously, therefore due to the anonymity of the data at source, once completed it is not possible for participants to withdraw their data from the study. However, you will be given the option to provide your name and contact details if you are happy

for us to contact you for further information based on your responses. Those who provide these details have the option to withdraw from the study up to 7 days after submission, by contacting Sophie Clohessy,.

6. Each individual is advised to fully consider, with others if necessary and prior to participation, any disadvantages, side effects, risks and/or discomforts that may arise from participation in this research.

7. Unless specifically agreed otherwise, all information will be held as confidential and will not be distributed outside of the research team.

8. Your data may be used as a source for future research, including research work for publication.

This research has been favourably reviewed by the University's Biomedical and Scientific Research Ethics Committee, Approval Reference: BSREC 111/20-21 dated: 17/01/2022. Dissatisfaction with the conduct of this research may be referred to the person below, who is a senior University of Warwick official entirely independent of this study: Head of Research Governance, Research & Impact Services, University House, University of Warwick, Coventry, CV4 8UW; Tel: 024 76 75733;

X→

consentform **Before you continue please understand that you are confirming your agreement with the following:**

○ I give my consent to my data submitted within this questionnaire being used for the purposes stated above including being happy for my data to be used in future research (1)

○ I do not consent, and I do not wish to take part (2)

*Skip To: End of Survey If Before you continue please understand that you are confirming your agreement with the following: = I do not consent, and I do not wish to take part*

Page Break

End of Block: information sheet

Start of Block: Work in West Midlands

Age **Are you aged 18 or over?**

◯ Yes (1)

◯ No (2)

*Display This Question:*

*If Are you aged 18 or over? = No*

End of survey **Thank you for your interest in our survey. Unfortunately we are only able to accept responses from people who are aged 18 or over.**

**Please click on the next page to end the survey.**

*Skip To: End of Survey If Thank you for your interest in our survey. Unfortunately we are only able to accept responses fro... Is Displayed*

Page Break

County **Which county in the Clinical Research Network West Midlands region is your employing organisation based within?**

▼ Shropshire (1) ... I don't work in any of these counties (7)

*Display This Question:*

*If Which county in the Clinical Research Network West Midlands region is your employing organisation... = I don't work in any of these counties*

End of survey **Thank you for your interest in our survey. Unfortunately we are only interested in responses from people who work within the Clinical Research Network West Midlands region.**

**Please click on the next page to end the survey.**

*Skip To: End of Survey If Thank you for your interest in our survey. Unfortunately we are only interested in responses from... Is Displayed*

**End of Block: Work in West Midlands**

**Start of Block: Current use of digital tools**

Use of tools
Please think about the clinical, health or social care research that you are currently working on or have worked on in the last two years. We are interested to learn whether you have used any **digital tools** during any stage of your research studies.

In this study a **digital tool** is defined as an alternative to paper based methods that is IT based or an online platform that aids any aspect of the research study (stages of clinical research include but are not limited to; set up of research studies, participant recruitment, consent, participant retention, intervention delivery, data collection, data analysis, monitoring, patient reported outcomes).

Examples of digital tools might include but are not limited to: Electronic databases for participant screening Social media platforms for participant recruitment Communication within your research team (e.g., reporting adverse events via text message or email) Electronic consent Text messaging for participant retention Digital platforms for collecting survey data

**Thinking about the clinical, health or social care research studies you are currently working on or have worked over the past two years, have you used digital tools to assist in the operation, management or coordination of these studies?**

- Yes (1)
- No (2)
- I have delegated digital tool use to colleagues (3)

---

*Display This Question:*

*If Please think about the clinical, health or social care research that you are currently working on... = I have delegated digital tool use to colleagues*

Q238 **Thank you for your interest in our survey. Please forward the link of this survey to a relevant team member.**

*Skip To: End of Survey If Thank you for your interest in our survey. Please forward the link of this survey to a relevant t... Is Displayed*

---

*Display This Question:*

*If Please think about the clinical, health or social care research that you are currently working on... = No*

X→

If not used digital

**What barriers have you encountered that have prevented you using digital tools in your clinical, health or social care research? Please select all answer/s that apply.**

- ▢ Lack of budget (1)
- ▢ Stick to what worked in the past (2)
- ▢ I wasn't aware of digital tools I could use (3)
- ▢ I would need training to use digital tools (4)
- ▢ I was concerned this would bias participant recruitment (5)
- ▢ Other (6) ________________________________________________

*Display This Question:*

*If Please think about the clinical, health or social care research that you are currently working on... = No*

Intentions

**Please rate the following statement on a 1 - 5 scale (1 "strongly disagree") and (5 "strongly agree")**

**"I intend to use digital tools in future clinical, health or social care research studies"**

○ 1 Strongly disagree (1)

○ 2 (2)

○ 3 (3)

○ 4 (4)

○ 5 Strongly agree (5)

Page Break

*Display This Question:*

*If Please think about the clinical, health or social care research that you are currently working on... = No*

Opt in non digital **By taking part in this survey, you also have the opportunity to take part in a qualitative one to one interview (most likely conducted remotely and at a time convenient for you). We anticipate the interview will take approximately between 30 and 60 minutes.**

**During this interview, we will explore reasons for not using digital tools in your clinical, health or social care research in further depth and your views towards these tools.**

**If you are interested in taking part, please provide your name and email address below and we will be in touch shortly to provide further details.**

○ Yes I am interested in providing my contact details (1)

○ No, I opt out of taking part in a qualitative interview (2)

---

*Display This Question:*

*If By taking part in this survey, you also have the opportunity to take part in a qualitative one to... = No, I opt out of taking part in a qualitative interview*

End of survey **Please click on the next page to complete the survey.**

*Skip To: End of Survey If Please click on the next page to complete the survey. Is Displayed*

---

*Display This Question:*

*If By taking part in this survey, you also have the opportunity to take part in a qualitative one to... = Yes I am interested in providing my contact details*

Full name **Full Name**

________________________________________________________________

---

*Display This Question:*

*If By taking part in this survey, you also have the opportunity to take part in a qualitative one to... = Yes I am interested in providing my contact details*

Email address **Email Address**

________________________________________________________________

---

*Display This Question:*

*If By taking part in this survey, you also have the opportunity to take part in a qualitative one to... = Yes I am interested in providing my contact details*

Job title **Job Title**

________________________________________________________________

---

*Display This Question:*

*If By taking part in this survey, you also have the opportunity to take part in a qualitative one to... = Yes I am interested in providing my contact details*

Organisation **Which organisation do you work for?**

________________________________________________________________

---

*Display This Question:*

*If By taking part in this survey, you also have the opportunity to take part in a qualitative one to... = Yes I am interested in providing my contact details*

complete survey **Please click on the next page to complete the survey.**

*Skip To: End of Survey If Please click on the next page to complete the survey. Is Displayed*

**End of Block: Current use of digital tools**

---

**Start of Block: Introduction**

Instructions **On the next page we will ask you some questions about you.**

**End of Block: Introduction**

---

**Start of Block: Demographics**

General role **What is/was your role in the most <u>recent</u> clinical, health or social care research you were involved with?**

- ◯ Chief Investigator (3)
- ◯ Clinical Researcher (1)
- ◯ Data Analyst (9)
- ◯ Principal Investigator (2)
- ◯ Research Manager (7)
- ◯ Research Nurse (8)
- ◯ Trial/Study Coordinator (4)
- ◯ Trial/Study Data Manager (6)
- ◯ Trial/Study Manager (5)
- ◯ Other (please provide further details below) (1) ________________________________________________

---

X→

Organisation **What organisation do you work for?**

- ◯ Clinical Trials Unit (2)
- ◯ NHS Trust (4)
- ◯ Primary Care (3)
- ◯ University (1)
- ◯ Other (please provide further details below) (5) ________________________________________________

---

Place of work **Please provide the specific place of work (optional)**

________________________________________________________________

**End of Block: Demographics**

**Start of Block: All examples**

Examples **Please provide a list of digital tools you are currently using or have used in your clinical, health or social care research studies over the last two years (including those still running). You can provide as many examples as you like.**

Examples **List of digital tools used in the last two years**

Page Break

Instructions **We will now ask you some more in depth questions about <u>two</u> of the digital tools you provided.**

**Please choose one digital tool which you believe <u>worked MOST effectively</u> and one you believe <u>worked LEAST effectively,</u> in terms of operating/managing/coordinating the research study.**

**Please consider your examples based on any clinical/health/social research projects you have worked on in the last 2 years (including those still running)**

**We will ask you a block of questions about each example. On the next page, we will ask you about your first example.**

**End of Block: All examples**

---

**Start of Block: Least effective tool**

Intro We will now ask you a block of questions about a digital tool you have used that has worked LEAST EFFECTIVELY in terms of operating, managing or coordinating a clinical/health/social research study.

Please consider your answer based on any clinical/health/social research projects you have worked on in the last 2 years (including those still running)

Page Break

Least effective eg

**Please state which digital tool you have used that worked LEAST EFFECTIVELY in terms of operating/managing/coordinating a clinical/health/social research study?**

**Please provide a very brief explanation of the tool/method.**

**(You can additionally provide a website link to a publication or trial website.)**

________________________________

________________________________

________________________________

________________________________

________________________________

Page Break

Least eff stage
**This an example of a digital tool relating to...**
(please select your answer using the drop down box)

▼ Data analysis (1) ... Other (10)

Least eff effective **Relative to your experience of other digital tools used in clinical research, how ineffective do you rate this tool?**

**0 = no less effective than other tools, 10 = much less effective than other tools.**

| | 0 | 1 | 2 | 3 | 4 | 5 | 6 | 7 | 8 | 9 | 10 |
|---|---|---|---|---|---|---|---|---|---|---|---|
| 9 () | | | | | | | | | | | |

Least eff details **In what way is/was the tool/platform ineffective?**

________________________________________________________________

________________________________________________________________

________________________________________________________________

________________________________________________________________

________________________________________________________________

Least advantages **Based on your experience, please list any advantages that the use of this digital tool has brought to your clinical, health or social care research (If applicable).**

________________________________________________________________

________________________________________________________________

________________________________________________________________

Least disadvantages
**Based on your experience, please list any disadvantages that the use of this digital tool has brought to your clinical, health or social care research (If applicable).**

________________________________________________________________

________________________________________________________________

________________________________________________________________

________________________________________________________________

________________________________________________________________

Least eff status **You have provided a digital tool from a project you are currently working on or previously worked on. What is the status of this project?**

- ○ Setting up (1)
- ○ Ongoing (2)
- ○ Completed (3)

Study managing **What is/was the study speciality area?**
(please select your answer using the drop down box)

▼ Ageing (1) ... Other (31)

Least eff study type
**This digital tool was used in ...**

○ A Clinical Trial (2)

○ Clinical Research (1)

○ Health Research (3)

○ Social Care Research (4)

○ Other (5) ________________________________________________

---

Least eff user **Who is/was the end user of this digital tool? (Please tick all that apply)**

▢ Carer/family member of research participant (6)

▢ Front line health and care staff (1)

▢ Principal Investigators (3)

▢ Research delivery workforce (2)

▢ Research participants (4)

▢ Other/Multiple users (Further information can be added in the text box) (5)

________________________________________________

---

Page Break

Least eff goal **What is/was the main goal of this digital tool? (Please tick all that apply)**

- ☐ Deal with Covid related restrictions (3)
- ☐ Facilitate good data management / Information governance (6)
- ☐ Increase patient accessibility (2)
- ☐ Improve data handling for analysis/modelling (5)
- ☐ Reduce time/Increase efficiency (1)
- ☐ Other (7) ________________________________________________

---

Previous method repl **What previous (i.e., non-digital/paper-based) method did this digital tool replace?**

________________________________________________________________

---

Least novelty **Is this digital tool novel/bespoke to your project team or used widely?**

- ○ Novel/bespoke to my project team (1)
- ○ Novel/bespoke to my organisation (2)
- ○ Used widely (3)

---

Page Break

least eff implement **How essential was this tool to successfully implement the research (relative to alternative non-digital tools)?**

- ○ Highly non-essential 1 (1)
- ○ Somewhat non-essential 2 (2)
- ○ Somewhat essential 3 (3)
- ○ Highly essential 4 (4)

Page Break

Least eff integ **How effective was/is this tool at integrating with other healthcare or research systems used during your research?**

- Not applicable 1 (1)
- Highly ineffective 2 (2)
- Somewhat ineffective 3 (3)
- Somewhat effective 4 (4)
- Highly effective 5 (5)

Page Break

Least eff tech lit **In your opinion, what level of technical literacy is/was required to use or access this digital tool?**

- Very difficult to use/access 1  (1)
- Somewhat difficult to use/access 2  (2)
- Somewhat easy to use/access 3  (3)
- Very easy to use/access 4  (4)

Page Break

Least eff training **Did you require training in order to use this digital tool?**

- Yes (1)
- No (2)

Page Break

Least eff costs **What costs were associated with acquiring this tool? (Please tick all that apply)**

- ▢ A one-off up-front fee (e.g. outright purchase) (1)
- ▢ Periodic recurring fee (e.g. monthly/annual license/maintenance fee) (2)
- ▢ No fee (e.g. open source) (3)
- ▢ Other (4) ________________________________________________
- ▢ I'm not sure (5)

Least eff comments **Please provide any further comments or information in the text box below about your digital tool example (optional)**

________________________________________________________________

________________________________________________________________

________________________________________________________________

________________________________________________________________

________________________________________________________________

**End of Block: Least effective tool**

**Start of Block: Most effective tool**

Intro We will now ask you a block of questions about a digital tool you have used that worked MOST EFFECTIVELY in terms of operating, managing or coordinating a clinical/health/social research study.

Please consider your answer based on any clinical/health/social research projects you have worked on in the last 2 years (including those still running)

Page Break

Most effect e.g

**Please state which digital tool you have used worked MOST EFFECTIVELY in terms of operating/managing/coordinating clinical/health/social research?**

**Please provide a very brief explanation of the tool/method.**

**(You can additionally provide a website link to a publication or trial website.)**

________________________________

________________________________

________________________________

________________________________

________________________________

Page Break

Most eff stage
**This an example of a digital tool relating to...**
(please select your answer using the drop down box)

▼ Data analysis (1) ... Other (10)

Most eff effective **Relative to your experience of other digital tools used in clinical research, how effective do you rate this tool?**

**0 = no more effective than other tools, 10 = much more effective than other tools.**

| | 0 | 1 | 2 | 3 | 4 | 5 | 6 | 7 | 8 | 9 | 10 |
|---|---|---|---|---|---|---|---|---|---|---|---|
| 9 () | | | | | | | | | | | |

Most eff details **In what way is/was the tool/platform effective?**

________________________________

________________________________

________________________________

________________________________

________________________________

Most eff advantages **Based on your experience, please list any <u>advantages</u> that the use of this digital tool has brought to your clinical, health or social care research (If applicable).**

________________________________

________________________________

________________________________

Most eff disadvantag
**Based on your experience, please list any disadvantages that the use of this digital tool has brought to your clinical, health or social care research (If applicable)**

________________________________________________________________

________________________________________________________________

________________________________________________________________

________________________________________________________________

________________________________________________________________

Most eff status **You have provided a digital tool from a project you have worked/currently working on. What is the status of this project?**

○ Setting up (1)

○ Ongoing (2)

○ Completed (3)

Most eff speciality **What is/was the study speciality area?**
(please select your answer using the drop down box)

▼ Ageing (1) ... Other (31)

Most eff
**This digital tool was used in ...**

- ○ A Clinical Trial (2)
- ○ Clinical Research (1)
- ○ Health Research (3)
- ○ Social Care (4)
- ○ Other (5) ________________________________________________

---

X→

Most eff user **Who is/was the end user of this digital tool? (Please tick all that apply)**

- ▢ Carer/family member of research participant (6)
- ▢ Front line health and care staff (1)
- ▢ Principal Investigators (3)
- ▢ Research delivery workforce (2)
- ▢ Research participants (4)
- ▢ Other/Multiple users (Further information can be added in the text box) (5) ________________________________________________

---

Page Break

Most eff end goal **What is/was the main goal of this digital tool? (Please tick all that apply)**

- ☐ Deal with Covid related restrictions (3)
- ☐ Facilitate good data management / Information governance (6)
- ☐ Increase patient accessibility (2)
- ☐ Improve data handling for analysis/modelling (5)
- ☐ Reduce time/Increase efficiency (1)
- ☐ Other (7) ________________________________________________

most eff previous **What previous (i.e., non-digital/paper-based) method did this digital tool replace?**

________________________________________________________________

Most eff novelty **Is this digital tool novel/bespoke to your project team or used widely?**

- ○ Novel/bespoke to my project team (1)
- ○ Novel/bespoke to my organisation (2)
- ○ Used widely (3)

Page Break

most eff implement **How essential was this tool to successfully implement the research (relative to alternative non-digital tools)?**

- Highly non-essential 1 (1)
- Somewhat non-essential 2 (2)
- Somewhat essential 3 (3)
- Highly essential 4 (4)

Page Break

Most eff integration **How effective was/is this tool at integrating with other healthcare or research systems used during your research?**

- Not applicable 1 (1)
- Highly ineffective 2 (2)
- Somewhat ineffective 3 (3)
- Somewhat effective 4 (4)
- Highly effective 5 (5)

Page Break

Most eff tech lit **In your opinion, what level of technical literacy is/was required to use or access this digital tool?**

- Very difficult to use/access 1 (1)
- Somewhat difficult to use/access 2 (2)
- Somewhat easy to use/access 3 (3)
- Very easy to use/access 4 (4)

Page Break

Most eff training **Did you require training in order to use this digital tool?**

- Yes (1)
- No (2)

Page Break

Most eff costs **What costs were associated with acquiring this tool? (Please tick all that apply)**

- ▢ A one-off up-front fee (e.g. outright purchase) (1)
- ▢ Periodic recurring fee (e.g. monthly/annual license/maintenance fee) (2)
- ▢ No fee (e.g. open source) (3)
- ▢ Other (4) ________________________________________________
- ▢ I'm not sure (5)

---

Most eff comments **Please provide any further comments or information in the text box below about your digital tool example (optional)**

________________________________________________________________

________________________________________________________________

________________________________________________________________

________________________________________________________________

________________________________________________________________

**End of Block: Most effective tool**

---

**Start of Block: Technology Anxiety**

Tech anxiety
**Technology Anxiety**

**Think about the digital tool example you provided. Please rate the following statements**

**on a "Strongly Disagree" to "Strongly Agree" scale.**

| | **Strongly Disagree** (1) | **Very Much Disagree** (2) | **Disagree** (4) | **Not Sure** (5) | **Agree** (6) | **Very Much Agree** (7) | **Strongly Agree** (8) |
|---|---|---|---|---|---|---|---|
| **I like to keep up with new technology** (11) | ○ | ○ | ○ | ○ | ○ | ○ | ○ |
| **I am open to change my work habits if necessary** (20) | ○ | ○ | ○ | ○ | ○ | ○ | ○ |
| **I am likely to become technology-oriented due to the nature of my field** (21) | ○ | ○ | ○ | ○ | ○ | ○ | ○ |
| **If I were given enough time and sufficient resources to absorb change, I am likely to accept it** (22) | ○ | ○ | ○ | ○ | ○ | ○ | ○ |
| **Overall, I am not that anxious about using technology** (23) | ○ | ○ | ○ | ○ | ○ | ○ | ○ |

**End of Block: Technology Anxiety**

Start of Block: Opt in interviews

Qualitative survey **By taking part in this survey, you also have the opportunity to take part in a qualitative one to one interview (most likely conducted remotely and at a time convenient for you). We anticipate the interview will take between 30-60 minutes.**

**During this interview, we will explore your experiences about using digital tools in clinical research in further depth. We will ask you questions about the information you provided in this survey.**

**This study is part of a wider project to learn from people who have used digital tools within their research, learn about the benefits and drawbacks of these methods and inform other researchers about the digital tools available to them.**

**If you are interested in taking part, please provide your name and email address below and we will be in touch shortly to provide further details.**

○ Yes I am interested in providing my contact details and I consent to the answers on this survey being discussed during the interview (1)

○ No, I opt out of taking part in a qualitative interview (2)

---

*Display This Question:*

*If By taking part in this survey, you also have the opportunity to take part in a qualitative one to... = Yes I am interested in providing my contact details and I consent to the answers on this survey being discussed during the interview*

Full name **Full Name**

________________________________________________________________

---

*Display This Question:*

*If By taking part in this survey, you also have the opportunity to take part in a qualitative one to... = Yes I am interested in providing my contact details and I consent to the answers on this survey being discussed during the interview*

Email address **Email Address**

________________________________________________________________

*Display This Question:*

*If By taking part in this survey, you also have the opportunity to take part in a qualitative one to... = Yes I am interested in providing my contact details and I consent to the answers on this survey being discussed during the interview*

Job role **Job Title**

________________________________________________________________

*Display This Question:*

*If By taking part in this survey, you also have the opportunity to take part in a qualitative one to... = Yes I am interested in providing my contact details and I consent to the answers on this survey being discussed during the interview*

Name of organisation **Which organisation do you work for?**

________________________________________________________________

**End of Block: Opt in interviews**

**Start of Block: Free text box**

Further Information **Please use this text box to provide any further information that you think may be relevant about your experience of using digital tools in your clinical research.**

________________________________________________________________

________________________________________________________________

________________________________________________________________

________________________________________________________________

________________________________________________________________

comments **Please leave any comments about the study below. In particular, please tell us if there was anything unclear or confusing about the study or any questions that you think we should include next time.**

________________________________________________________________

________________________________________________________________

________________________________________________________________

________________________________________________________________

________________________________________________________________

**End of Block: Free text box**

---

**Start of Block: Debriefing sheet**

Debriefing sheet
**Debriefing sheet**

**Study title: Digitising Clinical Research**

Thank you for taking part in the questionnaire-based study. Researchers Sophie Clohessy1, Prof Theo Arvanitis1, Dr Carla Toro1, Dr Mark Elliott1 carried out this study supported by collaborators Mark Evans2,3 Carly Craddock2,3.

1) Institute of Digital Health care, WMG, University of Warwick 2) The Royal Wolverhampton NHS Trust, Wolverhampton, UK, 3) National Institute for Health Research Clinical Research Network West Midlands, Birmingham, UK).

The purpose of this study is to identify local examples of digital platforms and approaches along with any barriers that may have delayed or stopped use of any digital tools. By taking part in this study, you had the opportunity to provide your contact details. If you chose to do so, we may invite you to take part in a qualitative interview study which will ask you about your experience with digital tools/platforms in further depth. Please be reminded that all data collected will remain confidential and anonymised. As mentioned in the information sheet, please note if you only partially completed the survey, we will still use the information that you provided up to the point that you stopped the survey. The findings of this study will be written up and published in a peer reviewed journal. The data will be stored securely on University of Warwick secure shared servers (M drive) for the standard data retention period of 10 years. The only people able to access the data will be the investigators of the project; Researchers Sophie Clohessy, Professor Theo Arvanitis, Dr Carla Toro, Dr Mark Elliott (Institute Digital Healthcare, WMG, University of Warwick).

If you have questions or concerns about the study please feel free to contact Sophie Clohessy,.

You may wish to keep a copy of this debriefing sheet for your records.

Thank you for taking your time to complete our study, we hope you enjoyed the experience.

**Please click on the next page to complete the study.**

**End of Block: Debriefing sheet**

---

### Supplementary Information 2b - Online Survey: R&D Department Staff

Start of Block: information sheet

Information sheet **PROJECT TITLE**: **Exploring Digital Approaches within R&D Departments**

**NAMES OF RESEARCHERS**: Sophie Clohessy1, Prof Theo Arvanitis1, Dr Carla Toro1, Dr Mark Elliott1 (1. Institute of Digital Health care, WMG, University of Warwick)

**NAMES OF COLLABORATORS:** Carly Craddock2,3, Mark Evans2,3 (2. The Royal Wolverhampton NHS Trust, Wolverhampton, UK, 3. National Institute for Health Research Clinical Research Network West Midlands, Birmingham, UK).

This sheet seeks to provide information, and advice, with respect to an individual's participation in support of the specified research project:

1. The study aims to investigate how online and digital tools/methods are being used for clinical, health and social care research. We are particularly interested to determine which approaches work well and which don't and use these findings to share best practice with the clinical, health and social care research community.

2. This study is funded by the National Institute of Health Research Clinical Research Network West Midlands (CRN West Midlands). The funding for this project has been awarded to Sophie Clohessy, Prof Theo Arvanitis, Dr Carla Toro, Dr Mark Elliott (University of Warwick). The project will be led by The University of Warwick.

3. Participation in this research is totally voluntary, and assurances are given to the effect that no negative consequences will arise from refusal to participate in the research project.

4. Your consent for your data to be used in this questionnaire will be gained by your ticking the consent question at the end of this page, so by ticking this box you agree that your submitted data can be used in the aforementioned study.

5. Your data will be collected anonymously, therefore due to the anonymity of the data at source, once completed it is not possible for participants to withdraw their data from the study. However, you will be given the option to provide your name and contact details if you are happy for us to contact you for further information based on your responses. Those who provide these details have the option to withdraw from the study up to 7 days after submission, by contacting

Sophie Clohessy,.

6. Each individual is advised to fully consider, with others if necessary and prior to participation, any disadvantages, side effects, risks and/or discomforts that may arise from participation in this research.

7. Unless specifically agreed otherwise, all information will be held as confidential and will not be distributed outside of the research team.

8. Your data may be used as a source for future research, including research work for publication.

This research has been favourably reviewed by the University's Biomedical and Scientific Research Ethics Committee, Approval Reference: BSREC 111/20-21 dated: 17/01/2022. Dissatisfaction with the conduct of this research may be referred to the person below, who is a senior University of Warwick official entirely independent of this study: Head of Research Governance, Research & Impact Services, University House, University of Warwick, Coventry, CV4 8UW; Tel: 024 76 75733;

---

X→

consentform **Before you continue please understand that you are confirming your agreement with the following:**

◯ I give my consent to my data submitted within this questionnaire being used for the purposes stated above including being happy for my data to be used in future research (1)

◯ I do not consent, and I do not wish to take part (2)

*Skip To: End of Survey If Before you continue please understand that you are confirming your agreement with the following: = I do not consent, and I do not wish to take part*

---

Page Break

End of Block: information sheet

---

Start of Block: Work in West Midlands

X→

County **Which county in the Clinical Research Network West Midlands is your employing organisation based within?**

▼ Shropshire (1) ... I don't work in any of these counties (7)

---

*Display This Question:*

*If Which county in the Clinical Research Network West Midlands is your employing organisation based... = I don't work in any of these counties*

End of survey **Thank you for your interest in our survey. Unfortunately we are only interested in responses from people who work within the Clinical Research Network West Midlands.**

**Please click on the next page to end the survey.**

*Skip To: End of Survey If Thank you for your interest in our survey. Unfortunately we are only interested in responses from... Is Displayed*

End of Block: Work in West Midlands

---

Start of Block: Current use of digital tools

X→

Use of tools
**Please think about the clinical, health or social care research that you are currently working on/have worked on in the last two years. We are interested to learn whether you have used any digital tools in the R & D department you work in. Specifically, we are interested in digital tools used for set up, recruitment of participants and management of clinical research studies.**

**In this study a digital tool is defined as an alternative to paper-based methods that is IT based or an online platform that aids any aspect of the research study set up, recruitment of participants and management.** Electronic screening for participants at site level Remote monitoring via smart phone applications Local social media routes R & D department's website Text alerts
**Think about the clinical research studies you have assisted with in your R & D**

**department over the past two years. Has your department used digital tools in the set up, recruitment and management/monitoring of clinical research?**

- ○ Yes (1)
- ○ No (2)

---

*Display This Question:*

*If Please think about the clinical, health or social care research that you are currently working on... = No*

If not used digital
**What barriers have you encountered that have prevented you using digital tools in your R & D department? Please select all answer/s that apply.**

- ☐ Concern digital tools would bias participant recruitment (8)
- ☐ GDPR concerns (10)
- ☐ Lack of budget (1)
- ☐ Stick to what worked in the past (2)
- ☐ Unaware of suitable digital tools (3)
- ☐ Would require training to use digital tools (7)
- ☐ Other (9) ________________________________________________

---

*Display This Question:*

*If Please think about the clinical, health or social care research that you are currently working on... = No*

Intentions

**Please rate the following statement on a 1 - 5 scale (1 "strongly disagree") and (5 "strongly agree")**

**"I intend to use digital tools for the set up, recruitment and management of future clinical, health or social care research studies"**

- ○ 1 Strongly disagree (1)
- ○ 2 (2)
- ○ 3 (3)
- ○ 4 (4)
- ○ 5 Strongly agree (5)

Page Break

*Display This Question:*

*If Please think about the clinical, health or social care research that you are currently working on... = No*

Opt in non digital **By taking part in this survey, you also have the opportunity to take part in a qualitative one to one interview (most likely conducted remotely and at a time convenient for you). We anticipate the interview will take approximately between 30 and 60 minutes.**

**During this interview, we will explore reasons for not using digital tools in set up, recruitment or monitoring of clinical, health or social care research in further depth and your views towards these tools.**

**If you are interested in taking part, please provide your name and email address below and we will be in touch shortly to provide further details.**

- ○ Yes I am interested in providing my contact details (1)
- ○ No, I opt out of taking part in a qualitative interview (2)

---

*Display This Question:*

*If By taking part in this survey, you also have the opportunity to take part in a qualitative one to... = Yes I am interested in providing my contact details*

Full name **Full Name**

________________________________________________________________

---

*Display This Question:*

*If By taking part in this survey, you also have the opportunity to take part in a qualitative one to... = Yes I am interested in providing my contact details*

Email address **Email Address**

________________________________________________________________

---

*Display This Question:*

*If By taking part in this survey, you also have the opportunity to take part in a qualitative one to... = Yes I am interested in providing my contact details*

Job title **Job Title**

________________________________________________________________

*Display This Question:*

*If By taking part in this survey, you also have the opportunity to take part in a qualitative one to... = Yes I am interested in providing my contact details*

Organisation **Which organisation do you work for?**

________________________________________________________________

*Display This Question:*

*If By taking part in this survey, you also have the opportunity to take part in a qualitative one to... = No, I opt out of taking part in a qualitative interview*

complete survey **Please click on the next page to complete the survey.**

*Skip To: End of Survey If Please click on the next page to complete the survey. Is Displayed*

**End of Block: Current use of digital tools**

---

**Start of Block: Introduction**

*Display This Question:*

*If Please think about the clinical, health or social care research that you are currently working on... = Yes*

Instructions **On the next page we will ask you some questions about you.**

**End of Block: Introduction**

---

**Start of Block: Demographics**

X→

Job role **What is your job role?**

- ○ Data Manager (11)
- ○ Director for Research (6)
- ○ Lead Research Nurse (9)
- ○ R&D Manager/Head of R&D (7)
- ○ Research and Development Facilitator (governance, study set-up) (8)
- ○ Research Delivery staff (10)
- ○ Other (please provide further details below) (5) ________________________________________________

---

Organisation **What organisation do you work for?**

- ○ Clinical trial unit (2)
- ○ Local Authority (6)
- ○ NHS Trust (4)
- ○ Primary Care (3)
- ○ University (1)
- ○ Other (please provide further details below) (5) ________________________________________________

---

Place of work **Please provide the specific place of work (optional)**

________________________________________________________________

**End of Block: Demographics**

Start of Block: Introduction to blocks

Intro to blocks **We are now going to ask you about digital tools used in R & D. A block of questions will be repeated for three different areas of clinical research studies: Set up Recruitment Management**

End of Block: Introduction to blocks

---

Start of Block: Set up

Intro set up **We will now ask you questions about digital tools used in R & D for the set up of clinical research studies.**

Page Break

Set up
**Has the Research and Development department you work in used any digital tools to set up studies, for example for feasibility, governance and authorisation (excluding Edge)?**

○ Yes (1)

○ No (2)

*Skip To: End of Block If Has the Research and Development department you work in used any digital tools to set up studies,... = No*

Set up examples **If Yes, please list the digital tool/s used by your R & D team to set up studies**

________________________________________________________________

________________________________________________________________

________________________________________________________________

________________________________________________________________

________________________________________________________________

Page Break

Intro set up **We will now ask you a series of questions about an example of a digital tool you just provided for set up of clinical research studies.**

**Please think about the tool you perceive to be the most effective. Please name the tool below**

________________________________________________________________

Hear Digital **How did you hear about this tool?**

- Approached by digital tool provider (1)
- Developed in house for specific need (2)
- Recommended by others (3)
- Saw advertised online/social media (4)
- Other (5) ________________________________________________

set up trainng **Did your R & D department require training in order to use this digital tool?**

- Yes (1)
- No (2)

*Display This Question:*

*If Did your R & D department require training in order to use this digital tool? = Yes*

training days **If yes, how many days training were required?**

- ◯ < 1 day (1)
- ◯ 1-5 days (2)
- ◯ > 5 days (3)

---

set up novel **Is this digital tool novel/bespoke to your project team or used widely?**

- ◯ Novel/bespoke to my project team (1)
- ◯ Novel/bespoke to my organisation (2)
- ◯ Used widely (3)

---

Page Break

Set up advantages **Based on your experience, please list any <u>advantages</u> of this digital tool? (If applicable)**

________________________________________________________________

________________________________________________________________

________________________________________________________________

________________________________________________________________

________________________________________________________________

Page Break

set up disadvantages
**Based on your experience, please list any disadvantages of this digital tool? (If applicable)**

________________________________________________________________

________________________________________________________________

________________________________________________________________

________________________________________________________________

________________________________________________________________

set up costs **What costs were associated with acquiring this tool?**

- ○ A one-off up-front fee (e.g. outright purchase) (1)
- ○ Periodic recurring fee (e.g. monthly/annual license/maintenance fee) (2)
- ○ No fee (e.g. open source) (3)
- ○ Other (4) ________________________________________________
- ○ I don't know the answer (5)

set up actual costs **Please state any details you have (and can disclose) on actual costs and any overall savings gained or expect to gain**

________________________________________________________________

________________________________________________________________

________________________________________________________________

________________________________________________________________

________________________________________________________________

Cost effective **How cost effective would you say this tool is in terms of whether it has added value/efficiency?**

- ○ Highly Ineffective 1 (1)
- ○ 2 (2)
- ○ 3 (3)
- ○ 4 (4)
- ○ 5 (5)
- ○ 6 (6)
- ○ Highly Effective 7 (7)

---

set up comments **Please provide any further comments or information in the text box below about your digital tool example. Please write n/a in the box if not applicable.**

________________________________________________________________

________________________________________________________________

________________________________________________________________

________________________________________________________________

________________________________________________________________

**End of Block: Set up**

---

**Start of Block: Recruitment**

Recruitment intro **We will now ask you questions about digital tools used in R & D for participant recruitment.**

---

Page Break

Recruitment
**Has the Research and Development department you work in used any digital means to recruit participants (please only include anything that is specific to your organisation and not anything that is mandated/provided by the study team and used study-wide)?**

○ Yes (1)

○ No (2)

*Skip To: End of Block If Has the Research and Development department you work in used any digital means to recruit partici... = No*

Recruitment example **If Yes, please list the digital tool/s used by your R & D team to recruit participants?**

________________________________________________________________

________________________________________________________________

________________________________________________________________

________________________________________________________________

________________________________________________________________

Page Break

Intro recruitment **We will now ask you a series of questions about an example of a digital tool you just provided for participant recruitment.**

**Please think about the tool you perceive to be the most effective. Please name the tool below**

________________________________________________________________

---

Recruitment hear **How did you hear about this tool?**

- ◯ Approached by digital tool provider (1)
- ◯ Developed in house for specific need (2)
- ◯ Recommended by others (3)
- ◯ Saw advertised online/social media (4)
- ◯ Other (5) ________________________________________________

---

Recruitment training **Did your R & D department require training in order to use this digital tool?**

- ◯ Yes (1)
- ◯ No (2)

---

*Display This Question:*

*If Did your R & D department require training in order to use this digital tool? = Yes*

Recruitment days **If yes, how many days training were required?**

- ○ < 1 day (1)
- ○ 1-5 days (2)
- ○ > 5 days (3)

---

Recruitment novel **Is this digital tool novel/bespoke to your project team or used widely?**

- ○ Novel/bespoke to my project team (1)
- ○ Novel/bespoke to my organisation (2)
- ○ Used widely (3)

---

Page Break

Recruitment advantag **Based on your experience, please list any advantages of this digital tool? (If applicable)**

________________________________________________________________

________________________________________________________________

________________________________________________________________

________________________________________________________________

________________________________________________________________

Page Break

Recruitment dis
**Based on your experience, please list any disadvantages of this digital tool? (If applicable)**

________________________________________________________________

________________________________________________________________

________________________________________________________________

________________________________________________________________

________________________________________________________________

Recruitment costs **What costs were associated with acquiring this tool?**

- ◯ A one-off up-front fee (e.g. outright purchase) (1)
- ◯ Periodic recurring fee (e.g. monthly/annual license/maintenance fee) (2)
- ◯ No fee (e.g. open source) (3)
- ◯ Other (4) ________________________________________________
- ◯ I don't know the answer (5)

cost effective **How cost effective would you say this tool is in terms of whether it has added value/efficiency?**

- ○ Highly Ineffective 1 (1)
- ○ 2 (2)
- ○ 3 (3)
- ○ 4 (4)
- ○ 5 (5)
- ○ 6 (6)
- ○ Highly Effective 7 (7)

---

Recruitment costs **Please state any details you have (and can disclose) on actual costs and any overall savings gained or expect to gain**

________________________________________________________________

________________________________________________________________

________________________________________________________________

________________________________________________________________

________________________________________________________________

---

Recruitment comments **Please provide any further comments or information in the text box below about your digital tool example. Please write n/a in the box if not applicable.**

________________________________________________________________

________________________________________________________________

________________________________________________________________

________________________________________________________________

End of Block: Recruitment

Start of Block: Management

Manage intro **We will now ask you questions about digital tools used in R & D for the management of clinical research studies.**

Page Break

X→

Manage **Has the Research and Development department you work in used any digital means to manage (i.e. progress, automate or alert for study procedures, study monitoring, financial management, study closure etc) a research study/studies (excluding Edge)?**

○ Yes (1)

○ No (2)

*Skip To: End of Block If Has the Research and Development department you work in used any digital means to manage (i.e. pr... = No*

Manage exmaple **If Yes, please tell us about the digital tool/s used by your R & D team to manage studies**

________________________________________________________________

________________________________________________________________

________________________________________________________________

________________________________________________________________

________________________________________________________________

Page Break

Manage intro We will now ask you a series of questions about an example of a digital tool you just provided for research study **management**.

Please think about the tool you perceive to be the most effective. Please name the tool below

________________________________________________________________

Manage hear **How did you hear about this tool?**

- ○ Approached by digital tool provider (1)
- ○ Developed in house for specific need (2)
- ○ Recommended by others (3)
- ○ Saw advertised online/social media (4)
- ○ Other (5) ________________________________________________

Manage training **Did your R & D department require training in order to use this digital tool?**

- ○ Yes (1)
- ○ No (2)

*Display This Question:*

*If Did your R & D department require training in order to use this digital tool? = Yes*

Manage days training **If yes, how many days training were required?**

◯ < 1 day (1)

◯ 1-5 days (2)

◯ > 5 days (3)

---

Manage novel **Is this digital tool novel/bespoke to your project team or used widely?**

◯ Novel/bespoke to my project team (1)

◯ Novel/bespoke to my organisation (2)

◯ Used widely (3)

---

Page Break

Manage advantage **Based on your experience, please list any advantages of this digital tool? (If applicable)**

________________________________

________________________________

________________________________

________________________________

________________________________

Page Break

Manage disadvantage
**Based on your experience, please list any disadvantages of this digital tool? (If applicable)**

________________________________________________________________

________________________________________________________________

________________________________________________________________

________________________________________________________________

________________________________________________________________

Manage costs **What costs were associated with acquiring this tool?**

- ○ A one-off up-front fee (e.g. outright purchase)  (1)
- ○ Periodic recurring fee (e.g. monthly/annual license/maintenance fee)  (2)
- ○ No fee (e.g. open source)  (3)
- ○ Other  (4) ________________________________________________
- ○ I don't know the answer  (5)

Cost effective **How cost effective would you say this tool is in terms of whether it has added value/efficiency?**

- Highly Ineffective 1 (1)
- 2 (2)
- 3 (3)
- 4 (4)
- 5 (5)
- 6 (6)
- Highly Effective 7 (7)

---

Manage costs **Please state any details you have (and can disclose) on actual costs and any overall savings gained or expect to gain**

________________________________________________________________

________________________________________________________________

________________________________________________________________

________________________________________________________________

________________________________________________________________

---

Manage further comme **Please provide any further comments or information in the text box below about your digital tool example. Please write n/a in the box if not applicable.**

________________________________________________________________

________________________________________________________________

________________________________________________________________

________________________________________________________________

__________________________________________________________________

End of Block: Management

Start of Block: Opt in interviews

X→

Qualitative survey **By taking part in this survey, you also have the opportunity to take part in a qualitative one to one interview (most likely conducted remotely and at a time convenient for you). We anticipate the interview will take between 30-60 minutes.**

**During this interview, we will explore your experiences about using digital tools in clinical research in further depth. We will ask you questions about the information you provided in this survey.**

**This study is part of a wider project to learn from people who have used digital tools within clinical research (including researchers) in order to learn about the benefits and drawbacks of these methods and inform researchers and related staff about the digital tools available to them.**

**If you are interested in taking part, please provide your name and email address below and we will be in touch shortly to provide further details.**

○ Yes I am interested in providing my contact details and I consent to the answers on this survey being discussed during the interview (1)

○ No, I opt out of taking part in a qualitative interview (2)

*Display This Question:*

*If By taking part in this survey, you also have the opportunity to take part in a qualitative one to... = Yes I am interested in providing my contact details and I consent to the answers on this survey being discussed during the interview*

Full name **Full Name**

__________________________________________________________________

*Display This Question:*

*If By taking part in this survey, you also have the opportunity to take part in a qualitative one to... = Yes I am interested in providing my contact details and I consent to the answers on this survey being discussed during the interview*

Email address **Email Address**

________________________________________________________________

*Display This Question:*

*If By taking part in this survey, you also have the opportunity to take part in a qualitative one to... = Yes I am interested in providing my contact details and I consent to the answers on this survey being discussed during the interview*

Job role **Job Title**

________________________________________________________________

*Display This Question:*

*If By taking part in this survey, you also have the opportunity to take part in a qualitative one to... = Yes I am interested in providing my contact details and I consent to the answers on this survey being discussed during the interview*

Name of organisation **Which organisation do you work for?**

________________________________________________________________

**End of Block: Opt in interviews**

---

**Start of Block: Free text box**

Further Information **Please use this text box to provide any further information that you think may be relevant about your experience of using digital tools in your role.**

________________________________________________________________

________________________________________________________________

________________________________________________________________

________________________________________________________________

________________________________________________________________

comments **Please leave any comments about the study below. In particular, please tell us if there was anything unclear or confusing about the study or any questions that you think we should include next time.**

________________________________________________________________

________________________________________________________________

________________________________________________________________

________________________________________________________________

________________________________________________________________

**End of Block: Free text box**

**Start of Block: Debriefing sheet**

Debriefing sheet
**Debriefing sheet**

**Study title: Digitising Clinical Research**

Thank you for taking part in the questionnaire-based study. Researchers Sophie Clohessy1, Prof Theo Arvanitis1, Dr Carla Toro1, Dr Mark Elliott1 carried out this study (1. Institute of Digital Health care, WMG, University of Warwick) with support from collaborators Carly Craddock2,3, Mark Evans2,3 (2. The Royal Wolverhampton NHS Trust, Wolverhampton, UK, 3. National Institute for Health Research Clinical Research Network West Midlands, Birmingham, UK).

The purpose of this study is to identify local examples of digital platforms and approaches along with any barriers that may have delayed or stopped use of any digital tools. By taking part in this study, you had the opportunity to provide your contact details. If you chose to do so, we may invite you to take part in a qualitative interview study which will ask you about your experience with digital tools/platforms in further depth. Please be reminded that all data collected will remain confidential and anonymised. As mentioned in the information sheet, please note if you only partially completed the survey, we will still use the information that you provided up to the point that you stopped the survey. The findings of this study will be written up and published in a peer reviewed journal. The data will be stored securely on University of Warwick secure shared servers (M drive) for the standard data retention period of 10 years. The only people able to access the data will be the investigators of the project; Researchers Sophie Clohessy, Dr Carla Toro, Dr Mark Elliott, Professor Theo Arvanitis (Institute Digital Healthcare, WMG, University of Warwick).

If you have questions or concerns about the study please feel free to contact Sophie Clohessy,.

You may wish to keep a copy of this debriefing sheet for your records.

Thank you for taking your time to complete our study, we hope you enjoyed the experience.

**Please click on the next page to complete the study.**

**End of Block: Debriefing sheet**
