## Supplementary File 3 for "Using Digital Tools in Clinical, Health and Social Care Research: A Mixed-Methods Study of UK Stakeholders"

### Supplementary 3a: Research staff– Digitising Research Qualitative Interview Questions

**Interview Introduction**

Research Fellow (RF) to thank participants for agreeing to participate in this research interview.

RF: Regarding the Participant Information Sheet and Consent Form we sent to you, did you have any questions about the study?

RF will give brief overview of the sections below before interview begins.

**Purpose of the study:**
To explore your experience of digital tools used in clinical research studies.

**Definition**:

- A digital tool can be used throughout different stages of clinical research.
- For example, set up, recruitment, participant retention, intervention delivery, data collection, data analysis and perhaps many more.
- Specific examples include; use of electronic health records to screen participants for eligibility for studies. Tools for recruitment of patients might include trial websites, social media, and email campaigns. Tools for data collection might include electronic platforms such as Prolific.

**Why are we doing this study?**

- This project is conducted by researchers at the Institute of Digital Healthcare, University of Warwick and funded by the NIHR CRN West Midlands.
- The use of digital tools has rapidly increased since the pandemic. Therefore, we are interested in exploring the experiences of stakeholders in the CRN West Midlands when using digital tools in their clinical research.
- We wish to know what has worked well, what not so well when using these tools. We wish to provide other researchers of local examples in the CRN West Midlands as well as recommendations/guidelines for future studies.

#### Interview Questions for Research staff

RF: I anticipate that the interview will take approximately 45 minutes to one hour. Please remember that we are interested in your experiences with digital tools in clinical research studies and there are no wrong or right answers. Please feel free to ask me to repeat questions if anything is unclear.

RF: We will now start recording the interview. RF to note time interview started recording.

For the purpose of the recording, I can confirm that consent has been taken from the participant and they had a chance to read the information sheet. We will now start the interview questions.

V2
17/08/2022

#### Section 1 - Opening question

**1.1. RF to ask participants if they have consented to providing their name, place of work, and organisation in report? If yes, participant to state those details now.**

**1.2. In a few sentences, please tell me about your role and a brief description of the main clinical research studies that you have worked on in the past two years (since 2020).**

#### Section 2 – Definition of digital tools

**2.1. In the context of clinical research, what does the term 'digital tool' mean to you?**
Prompt: How would you describe it to someone with no prior knowledge of the different digital tools used in clinical research?
Prompt: Please provide brief examples *they don't have to be a tool you have personally used.

#### Section 3- COVID-19 pandemic questions

**3.1. Please describe how/if COVID-19 pandemic affected your research?**
Prompt- Were the digital tools you provided as examples in the survey implemented due to Covid? Or other reasons? (If used before covid- why were they implemented).
Prompt-Utilise digital tools (or more digital tools) due to COVID instead of paper/non-digital tools?
Prompt- If yes, what were your thoughts on using more digital tools? (e.g., feel ready? Pressured)
Prompt- If yes, please describe the accelerated implementation of digital tools during the pandemic, how did this happen and were there any impacts of this?

#### Section 4 - Case study of MOST effective tool

**In the survey you provided an example of a digital tool which you deemed to be the most effective digital tool you have used.**
The example you provided was ………… and end user was …….. (Research Fellow to provide a sentence describing the tool/end user based on their survey answer)
**We would now like to ask you some questions about this digital tool and the clinical research study in which it was used**.

**4.1. Could you provide some background about the clinical research study in which the digital tool was used?**
Prompt: Project status? Study objective? Your role in this study?
Prompt: How was the digital tool used in the clinical research study? (Stage of project).

**4.2. Please describe the process in which this digital tool was chosen to be used in study? how and why was it chosen?**
Prompt: What previous tool/method did it replace? i.e., paper/non digital.
Prompt: Who chose the tool? (i.e., yourself, colleagues). Is there a process for deciding which tool is selected for use?
Prompt: Were other tools considered? If so, why was this tool chosen? E.g., commonly used?
Prompt: What criteria was used to decide whether it was a good tool or not? Was it helpful? How could this be improved?

Prompt: Do you think there should be a process that researchers could follow when choosing tools.

**4.4. You mentioned on the survey that training was required for this digital tool. Please describe what the training entailed? (Participants will only be asked this question if they said they had training for the digital tool example).**

Prompt- Training length? What parts were helpful? Could any areas be improved?

**4.5. You mentioned on the survey that you received no training for this digital tool (Participants will only be asked this question if they said no to training)**

Prompt- If no, why was training not provided? Do you believe it should have been provided? Would this have aided use/performance?

**4.6. We are interested in learning about the performance of this digital tool. Please tell me about your experience of using the digital tool?**

Research Fellow to immediately mention Prompt- why was this tool selected as most effective? ***Key follow up question** and Prompt- please consider the experience of using the digital tool to other tools (i.e., another digital tool or non-digital/paper-based method). ***Key follow up question.**

Prompt- did you find it easy or difficult to use the tool?
Prompt- what would you consider to be the benefits or drawbacks of this tool? (Particularly in relation to previous tools or non-digital/paper-based tool).
Prompt- did using the tool fulfil the goals of the research? (Please re-state the goals) If no, what alternative tools could have been used?
Prompt: Have you used this tool prior to this project? If yes, how long have you used this tool for?
Prompt- How many people in your team used the tool in the example you provided? (approx.).
Prompt- Can you describe the end user of this tool? Any formal feedback process?

**4.7. Please consider if there were any barriers to implementation/usage of this digital tool?**

Prompt- Barrier might be technological, logistical, cost, if participant facing-accessibility?
Prompt- If yes, was the barrier overcome? How was this achieved?

**4.8. Thinking of this digital tool, please describe any details you have (and can disclose) on actual costs and any overall savings gained or expect to gain?**

Prompt- is the tool cost effective?
If participants do not have information on exact costs…do you think that using digital tool makes savings on indirect costs i.e., adds efficiency? Saves time?

**4.9. We are interested in your future intentions to use this tool. Thinking of the current tool choice, would you do anything differently? Or utilise the tool again?**

Prompt- Will you continue to use the tool or similar in future projects? Or chose another tool?
Prompt- Why might this be? Please expand

#### Section 5 - Case study of LEAST effective tool

**5.1. In the survey you provided an example of a digital tool which you deemed to be the LEAST effective digital tool you have used.**
The example you provided was ………… and end user was …….. (Research Fellow to provide a sentence describing the tool/end user based on their survey answer)
**We would now like to ask you some questions about this digital tool and the clinical research study in which it was used**.

**5.2. Could you provide some background about the clinical research study in which the digital tool was used?**
Prompt: Project status? Study objective? Your role in this study?
Prompt: How was the digital tool used in the clinical research study? (Stage of project).

**5.3. Please describe the process in which this digital tool was chosen to be used in the study? how and why was it chosen?**
Prompt? Why it was thought to be effective when chosen but turned out not to be
Prompt: What previous tool/method did it replace? i.e., paper/non digital.
Prompt: Who chose the tool? (i.e., yourself, colleagues). Is there a process for deciding which tool is selected for use?
Prompt: Were other tools considered? If so, why was this tool chosen? E.g., commonly used?
Prompt: What criteria was used to decide whether it was a good tool or not? Was it helpful? How could this be improved?
Prompt: Do you think there should be a process that researchers could follow when choosing tools?

**5.4. a) You mention on the survey that training was required for this digital tool. Please describe what the training entailed?**

Prompt- Training length? What parts were helpful? Could any areas be improved?

**b) You mention on the survey that you received no training for this digital tool.**

Prompt- If no, why was training not provided? do you believe it should provided? Would this have aided use?/performance?

**5.5. We are interested in learning about the performance of this digital tool. Please tell me about your experience of using the digital tool and why it was deemed not effective?**
Prompt- why was this tool selected as least effective? ***Key follow up question**
Prompt- please consider the experience of using the digital tool to other tools (i.e., another digital tool or non-digital/paper-based method). ***Key follow up question.**

- Prompt- did you find it easy or difficult to use the tool?

Prompt- what would you consider to be the benefits or drawbacks of this tool? (Particularly in relation to previous tools or non-digital/paper-based tool).
Prompt- did using the tool fulfil the goals of the research? (Please re-state the goals) If no, what alternative tools could have been used?
Prompt: Have you used this tool prior to this project? If yes, how long have you used this tool for?
How many people in your team used the tool in the example you provided? (approx.).
Experience of end user? Any formal feedback process?

**5.6. Please consider if there were any barriers to implementation/usage of this digital tool?**
Prompt- Barrier might be technological, logistical, cost, if participant facing, accessibility?
Prompt- How was the barrier overcome? How was this achieved?

**5.7. Thinking of this digital tool, please describe any details you have (and can disclose) on actual costs and any overall savings gained or expect to gain?**

Prompt- is the tool cost effective?
If participants do not have information on exact costs… do you think that using digital tool makes savings on indirect costs i.e., adds efficiency? Saves time?

**5.8. We are interested in your future intentions to use this tool. Thinking of the current tool choice, would you do anything differently? Or utilise the tool again?**
Prompt- Are there any changes that could be made to make the tool more effective, if not what other tools could be used an alternative (if any)
Prompt- Will you continue to use the tool or similar in future projects? Or chose another tool?
Prompt- Why might this be? Please expand.

#### 6. General questions about digital tools

**6.1. Are there instances in your clinical research studies where digital tools could have been used instead of non-digital/paper-based tools?**
Prompt- If yes, why is this? which one? Would implementation of digital tools have affected the outcomes? In what way?

**6.2. Please consider if you have encountered any barriers that may have prevented you from using digital tools in your role?**
Prompt- Examples might include budget, training, just used what worked before, knowledge.

**6.4. When considering different digital tools in clinical research, what do you think would help researchers when deciding which digital tool to use and where might you find this advice?**
Prompt- What type of information might be useful? Case studies? Example where things have worked well/not so well? Database of tools? (Including costs, level of difficulty etc).

Prompt- What evidence of success would you look for when deciding whether to use a particular tool?
Prompt- Where would you expect to find advice? e.g., in house at local CRN or own in house research?

#### Final thoughts

**7.Is there anything I haven't asked you about that you think is important to add? Please tell me…**

**8. If I need to, would it be possible to come back to you if a further specific question arises as we conduct this research?**

**9.Thank you so much again for taking part, your time, knowledge and experience is greatly appreciated. Is there anything you would like to ask me about this research?**

RF to note time interview ended and stop recording.

### Supplementary 3b: Qualitative Interviews - R & D staff

**Interview Introduction**

Research Fellow (RF) to thank participants for agreeing to participate in this research interview.

RF: Regarding the Participant Information Sheet and Consent sheet we sent to you, did you have any questions about the study?

RF will give brief overview of the sections below before interview begins.

**Purpose of the study:**
To explore your experience of digital tools used in clinical research studies as an individual working in R & D.

**Definition**:
- A digital tool can be used throughout different stages of clinical research.
- We are interested in your experiences of using digital tools, specifically, we are interested in digital tools used for set up, recruitment of participants and management of clinical research studies.

**Why are we doing this study?**
- This project is conducted by researchers at the Institute of Digital Healthcare, University of Warwick and funded by the NIHR CRN West Midlands.
- The use of digital tools has rapidly increased since the pandemic. Therefore, we are interested in exploring the experiences of stakeholders in the CRN West Midlands when using digital tools in their clinical research.
- We wish to know what has worked well, what not so well when using these tools. We wish to provide other researchers and R & D departments local examples in the CRN West Midlands as well as recommendations/guidelines for future studies.

#### Interview Questions for R & D staff

RF: I anticipate that the interview will take approximately between 45 to 60 minutes. Please remember that we are interested in your experiences with digital tools in clinical research studies and there are no wrong or right answers. Please feel free to ask me to repeat questions if anything is unclear.

RF: We will now start recording the interview. RF to note time interview started recording.

For the purpose of the recording, I can confirm that consent has been taken from the participant and they had a chance to read the information sheet. We will now start the interview questions.

#### Section 1 - Opening question

**1.1. RF to ask participants if they have consented to providing their name, place of work, and organisation in report? If yes, participant to state those details now.**

#### Section 2 – Definition of digital tools

**2.1. In the context of clinical research, what does the term 'digital tool' mean to someone working in R & D?**

Prompt: How would you describe it to someone with no prior knowledge of the different digital tools used in clinical research?

Prompt: Please provide brief examples *they don't have to be tool you have personally used. Can you provide any examples of the tools someone working in an R & D department might encounter?

#### Section 3- COVID-19 pandemic questions

**In the survey you provided an example of an effective tool used in set up, recruitment, management of clinical research.**

**3.1. Please describe how/if COVID-19 pandemic affected your research, specifically with reference to your use of digital tools?**

Prompt- Were the digital tools you mentioned in the survey implemented due to Covid? Or other reasons? (If used before covid- why were they implemented).

Prompt-Utilise digital tools (or more digital tools) due to COVID instead of paper/non-digital tools?

Prompt- If yes, what were your thoughts on using more digital tools? (e.g., feel ready? Pressured)

Prompt- If yes, please describe the accelerated implementation of digital tools during the pandemic, how did this happen and were there any impacts of this?

#### Section 4 - Case study most effective tool used for set up

**In the survey you provided an example of a digital tool which you deemed to be the most effective digital tool you have used for the set-up of clinical research studies.**

**The example you provided was** ………… (Research Fellow to provide a sentence describing the tool based on their survey answer)

**We would now like to ask you some questions about this digital tool and the clinical research/trial in which it was used**.

**4.1. Could you provide some background about how the digital tool was used?**
Prompt: Briefly describe the tool and how was it used in a study/trial? Your role/how much did you interact with tool?

**4.2. Please describe the process in which this digital tool was chosen? how and why was it chosen?**
Prompt: What previous tool/method did it replace? i.e., paper/non digital.
Prompt: Who chose the tool? (i.e., yourself, colleagues). Is there a process for deciding which tool is selected for use?
Prompt: Were other tools considered? If so, why was this tool chosen? E.g., commonly used?
Prompt: What criteria was used to decide whether it was a good tool or not? Was it helpful? How could this be improved?
Prompt: Do you think there should be a process that researchers could follow when choosing tools.

**4.3. You mentioned on the survey that training <u>was</u> required for this digital tool. Please describe what the training entailed?**
Prompt- Training length? What parts were helpful? Could any areas be improved?

**4.4. You mentioned on the survey that you received <u>no</u> training for this digital tool.**
Prompt- If no, why was training not provided? Do you believe it should have been provided? Would this have aided use/performance?

**4.5. We are interested in learning about the performance of this digital tool. Please tell me about your experience of using the digital tool?**

Prompt- Why was this tool selected as most effective? ***Key follow up question**
Prompt- Please consider the experience of using the digital tool to other tools (i.e., another digital tool or non-digital/paper-based method). ***Key follow up question.**
Prompt- Did you find it easy or difficult to use the tool?
Prompt- What would you consider to be the benefits or drawbacks of this tool? (Particularly in relation to previous tools or non-digital/paper-based tool).
Prompt- Did using the tool fulfil the goals of the research? (Please re-state the goals) If no, what alternative tools could have been used?
Prompt: Have you used this tool prior to this project? If yes, how long have you used this tool for?
Prompt-How many people in your team used the tool in the example you provided? (approx.).
Can you describe the end user of this tool? Any formal feedback process?

**4.6. Please consider if there were any barriers to implementation/usage of this digital tool?**
Prompt- Barrier might be technological, logistical, cost, if participant facing-accessibility?
Prompt- If yes, was the barrier overcome? How was this achieved?

**4.7. Thinking of this digital tool, please describe any details you have (and can disclose) on actual costs and any overall savings gained or expect to gain?**
Prompt- Is the tool cost effective?
Prompt- If participants do not have information on exact costs… do you think that using digital tool makes savings on indirect costs i.e., adds efficiency? saves time?

**4.8. We are interested in your future intentions to use this tool. Thinking of the current tool choice, would you do anything differently? Or utilise the tool again?**
Prompt- Will you continue to use the tool or similar in future projects? Or chose another tool?
Prompt- Why might this be? Please expand.

#### Section 5- Case study most effective tool used for recruitment

#### Section 6- Case study most effective tool used for management

#### 7. General questions about digital tools

**7.1. Are there instances in your clinical research studies where digital tools could have been used instead of non-digital/paper-based tools?**
Prompt- If yes, why is this? which one? Would implementation of digital tools have affected the outcomes? In what way?

**7.2. Please consider if you have encountered any barriers that may have prevented you from using digital tools in your role?**
Prompt- Examples might include budget, training, just used what worked before, knowledge.

**7.3. When considering different digital tools in clinical research, what do you think would help researchers when deciding which digital tool to use and where might you find this advice?**
Prompt- What type of information might be useful? Case studies? Example where things have worked well/not so well? Database of tools? (Including costs, level of difficulty etc).
Prompt- What evidence of success would you look for when deciding whether to use a particular tool?
Prompt- Where would you expect to find advice? e.g., in house at local CRN or own in house research?

#### Final thoughts

**Is there anything I haven't asked you about that you think is important? Please tell me…**

**If I need to, would it be possible to come back to you if a further specific question arises as we conduct this research?**

**Thank you so much again for taking part, your time, knowledge and experience is greatly appreciated. Is there anything you would like to ask me about this research?**

RF to note time interview ended and stop recording.
